# Hyperinflammatory conditions, gender differences and mortality in Indian COVID-19 patients

**DOI:** 10.1101/2021.01.19.21250134

**Authors:** Fouzia Shoeb, Imran Hussain, Gazala Afrin, Shagufta T. Mufti, Tabrez Jafar, Syed T. Raza, Farzana Mahdi

## Abstract

**Purpose:** Evidence suggests that COVID-19 induces hyperinflammatory conditions and causes relatively more deaths in males than females. The purpose of this study was to analyze gender differences associated with various hyperinflammatory conditions (HIC) and mortality in the Indian COVID-19 patients

**Methods:** This study was conducted at the Era’s Lucknow Medical College and Hospital (ELMCH), ERA University, which is located in the northern part of India. Starting from July 4, 2020 till December 3, 2020 a total of 2997 patients were treated at ELMCH. We randomly collected blood samples from 150 severe COVID-19 patients (required oxygen) between August 10 and September 15, 2020 for analyzing the following HIC and associated laboratory markers: hyperferritinaemia (serum ferritin), hematological dysfunctions (lymphocytopenia and neutrophil to lymphocyte ratio), cytokinaemia (C-reactive protein), coagulopathy (D-dimer), liver inflammation (aspartate aminotransferase), renal inflammation (blood urea and creatinine), and hyperglycemia (random blood glucose). The threshold values/cut off limits of these laboratory markers used for analyzing the risk of mortality in male and female COVID-19 patients were set according to the scale validated recently by Webb et al, (2020).

**Results:** In the above cohort of consecutively admitted COVID-19 patients, analysis of various HIC revealed hyperferritinaemia (odd ratio: 2.9, 95% CI 1.4-6.0), hematological dysfunctions (odd ratio: 2.10, 95% CI 1.0-4.2), hepatic inflammation (odd ratio: 2.0, 95% CI 0.52-7.40), and coagulopathy (odd ratio: 1.5, 95% CI 1.50, 95% CI 0.50-4.60) were more prevalent and sever in male COVID-19 patients. Approximately 86% male to 64% female COVID-19 patients developed lymphocytopenia. Regarding mortality, while hyperferritinaemia (odd ratio: 1.70, 95% CI 0.37-7.43) and cytokinaemia (odd ratio: 1.60, 95% CI 0.37 −7.30) were strongly associated with mortality in male COVID-19 patients, coagulopathy (odd ratio: 3.30, 95% CI 0.31-35), and hematological dysfunctions (odd ratio: 1.70, 95% CI 0.27-10) were more commonly associated with mortality in female COVID-19 patients. Nearly 80% male and female COVID-19 patients, who died had developed ≥2 criteria of HIS criteria. Chronic renal disease was associated with more deaths in female than male COVID-19 patients (odd ratio: 2.0, 95% CI 0.54 - 7.4). While the mortality proportion was slightly higher in male (6.3%) than female (4.5%) COVID-19 patients, survival curves of the two genders were not different (hazard ratio: 1.02, 95% CI 0.71-1.40, P = 0. 953).

**Conclusion:** Distinct HIC were associated with the severity, and mortality in male and female COVID-19 patients. Coagulopathy and renal injury were detrimental, specifically, for female COVID-19 patients. The overall mortality proportion was around 5.3%. The above results suggest that gender differences associated with COVID-19 severity and mortality arise due to differences in various HIC. These results may help in developing personalized or gender based treatments for COVID-19 patients.

## Introduction

Coronavirus diseases-2019 (COVID-19) is a highly contagious disease characterized by respiratory failure and death in severe conditions (1). It is caused by a novel strain of coronavirus called severe acute respiratory syndrome corona virus 2 (SARS-CoV-2). COVID-19 has been found associated with various inflammatory conditions including macrophage activation, hematological dysfunctions, cytokinaemia or cytokine storm characterized by increased production of C-reactive protein (CRP), interleukins, and tumor necrosis factors-α (2–4) Coagulopathy, as identified by increased D-dimer formation, is another detrimental factor that has been reported to severely increase the risk for mortality in COVID-19 patients (5). Renal and liver inflammation has also been reported in severe COVID-19 cases (6-8).

Epidemiological data suggests that COVID-19 is mounting high mortalities in older people, specially, those of age ≥ 50 years (9,10). The risk of mortality increases further in patients having pre-existing morbidities such as hypertension, diabetes, renal disease, and respiratory complications (9,10). However, development of hyperinflammatory syndrome has been suggested as the potential cause of death in a significant proportion of COVID-19 patients (11). There is a pressing need for standardizing various clinical criteria and laboratory parameters for predicting the risk of severity and mortality associated with COVID-19. A recent cohort study has defined and validated various criteria for COVID-19 associated hyperinflammatory syndrome (11), which applied for predicting the risk of severity and mortality in COVID-19 patients. It has been suggested that patients who develop two or more criteria of hyperinflammatory syndrome most likely will progress to mechanical ventilation and deaths (11).

The purpose of the present study was to analyze hyperinflammatory conditions (HIC), and gender differences associated with mortality in Indian COVID-19 patients.

### Patients

This study was conduced at Era’s Lucknow Medical College and Hospital (ELMCH), which is located in Lucknow, a capital city of Utter Pradesh state of north India. ELMCH started receiving COVID-19 patients from 4 July 2020 onward and as of December 3, 2020 a total of 2997 patients were admitted to ELMCH. We randomly collected blood samples from COVID-19 patients on daily basis from 9 am to 2 pm between August 10, and September 15, 2020. A total of 150 (100 male and 50 females) samples were collected and used for measuring various laboratory markers associated with HIC. We also analyzed overall mortality in all COVID-19 patients (total 2997) admitted to ELMCH between July 4 and December 3, 2020. Body temperature measurement and RT-PCR method were used for COVID-19 diagnosis. A written and informed consent was taken from each patient or from his/her family members before drawing the blood.

## Methods

### Measurement of laboratory markers

All the measurements were performed on a daily bases using fresh blood samples. Complete blood cell counts and hemoglobin estimation were performed on Sysmex XS-800i automated hematology analyzer. Plasma/serum concentrations of ferritin, CRP, IL-6, Urea, Creatinine, and AST were measured using commercial kits from Ortho Clinical Diagnostics, New Jersey, USA. D-dimer was measured using commercially available kit (Cat# DZ179A-K) from Diazyme Laboratories, Poway, CA, USA. All the measurements were performed according to the instructions provided by the respective manufacturers, which are available at https://www.orthoclinicaldiagnostics.com/en-us/home/, and https://www.diazyme.com.

### Statistical analyses

Prism (GraphPad) was used for performing various statistical analyses. Student’s *t* test and/or ANOVA were used to find differences between the groups. Pearson product moment correlation analysis was performed to find the relationship among various laboratory markers. Fisher’s exact probability test was used for calculating relative risk/risk ratio, odd ratio, and sensitivity. Kaplan-Meier method was used for analyzing the survival curve and the significance was calculated using log-rank (Mantel-Cox) test. A P < 0.05 was considered significant.

## Results

A detailed analysis of various laboratory markers and their cut-off values including the minimum and maximum have been given in table 1. To find gender differences, we performed statistical analysis of various laboratory markers in male and female COVID-19 patients separately. Although the values of several laboratory markers were elevated above the normal in both male and female COVID-19 patients, significant gender differences (indicated by bold number) were observed for serum ferritin, lymphocyte counts, and neutrophil to lymphocyte ratio (N/L ratio).

**Table 1.** Analysis of laboratory markers associated with various hyperinflammatory conditions (HIC) in COVID-19 patients**\***

| Laboratory markers/parameters | Male | Female | AUROC (95% CI) | P |
| --- | --- | --- | --- | --- |
| Number of cases | 100 | 50 |  |  |
| Age (years) | 28 - 81 | 23 - 93 | 0.56 (0.47-0.68) | 0.1480 |
| Serum ferritin (µg/L) | **57 - 2720** | **13 - 1050** | **0.71 (0.62-0.79)** | **<0.001** |
| C-reactive protein (mg/dL) | 4 - 32 | 03 - 24.5 | 0.55 (0.45-0.65) | 0.3600 |
| D-dimer (µg/mL) | 0.43 - 5.73 | 0.54 - 4.30 | 0.57 (0.41-0.72) | 0.4170 |
| Aspartate aminotransferase (U/L) | 17 - 180 | 18 - 185 | 0.59 (0.50-0.69) | 0.0620 |
| White blood cell counts (10<sup>3</sup> per mm<sup>3</sup>) | 2.8 - 13.7 | 4.2 - 14 | 0.50 (0.40-0.60) | 0.9853 |
| Neutrophils (10<sup>3</sup> per mm<sup>3</sup>) | **5.9 - 9.8** | **5.0 - 9.6** | **0.62 (0.52-0.71)** | **0.0210** |
| Lymphocytes (10<sup>3</sup> per mm<sup>3</sup>) | **0.2 - 3.0** | **0.2 - 4.1** | **0.63 (0.53-0.72)** | **0.0120** |
| Neutrophil to lymphocyte ratio | **02 - 26** | **1.2 - 21** | **0.63 (0.53-0.72)** | **0.0110** |
| Hemoglobin (g/dL) | 6.8 - 15.8 | 6.2 - 13.5 | 0.69 (0.61-0.78) | <0.001 |
| Blood urea (mg/dL) | 18 - 240 | 11 - 248 | 0.55 (0.44-0.65) | 0.3541 |
| Creatinine (mg/dL) | 0.6 - 11.6 | 0.6 - 9.5 | 0.60 (0.49-0.69) | 0.0777 |
| Random blood glucose (mg/dL) | 38 - 500 | 41 - 464 | 0.53 (0.40-0.66) | 0.6596 |

\*Values are given as minimum and maximum, the bold numbers indicate significance of difference.

For establishing various HIC, we used the same cut-off values of the associated laboratory markers as validated recently by Webb et al, (2020) (11). These values are: ferritin ≥700 μg/L for hyperferritinaemia, CRP ≥15 mg/dL for cytokinaemia, neutrophil to lymphocyte ratio ≥10 for hematological dysfunctions, D-dimer ≥1.0 μg/mL for coagulopathy, AST ≥100 U/L for liver injury/inflammation (Table 2). We used the values of urea ≥ 70 mg/dL and creatinine ≥ 1.5 mg/dL for renal injury/inflammation, and random blood glucose (RBG) ≥ 200 mg/dL for hyperglycemia. Although Webb et al (2020) used a D-dimer concentration of 1.5 μg/mL for coagulopathy; they also suggested that a D-dimer concentration ≥1.0 μg/mL could be appropriate for predicting the risk of severity and mortality in COVID-19 patients. We also found that D-dimer concentration ≥1.0 was strongly associated with mortality in several COVID-19 patients who perished. Next, we have discussed gender susceptibility to develop various HIC by taking the above threshold values into consideration, and the detail statistical analysis is presented in Table 2.

**Table 2.** Prevalence of various HIC in male and female COVID-19 patients

| HIC (↓) | Cut-off limit | Male | Female | Risk ratio<br>(95% CI) | Odd ratio<br>(95% CI) | Sensitivity<br>(95% CI) | Specificity<br>(95% CI) | AUROC<br>(95% CI) |
| --- | --- | --- | --- | --- | --- | --- | --- | --- |
| **Hyperferritinaemia** | (Ferritin ≥ 700 µg/L) | 55/100 (55%) | 15/51 (30%) | 1.9 (1.2-2.9) | 2.9 (1.4-6.0) | 0.79 (0.67-0.87) | 0.44 (0.33-0.56) | 0.66 (0.53-0.79) |
| **Hematological dysfunction** | (N/L ratio ≥ 10) | 68/96 (71%) | 27/51 (54%) | 1.3 (0.10-1.75) | 2.1 (1.0- 4.2) | 0.72 (0.61-0.80) | 0.45 (0.31-0.60) | 0.62 (0.50-0.75) |
| **Cytokinaemia** | (CRP ≥ 15 mg/dL) | 14/98 (14%) | 06/51 (12%) | 1.2 (0.49-2.91) | 1.2 (0.44-3.40) | 0.70 (0.46-0.88) | 0.34 (0.26-0.43) | 0.69 (0.47-0.90) |
| **Coagulopathy** | (D-dimer ≥ 1.0 µg/mL) | 24/37 (65%) | 11/20 (55%) | 1.2 (0.74-1.90) | 1.5 (0.50-4.60) | 0.69 (0.51-0.83) | 0.41 (0.21-0.64) | 0.63 (0.43-0.83) |
| **Hepatic inflammation** | (AST ≥ 100 U/L) | 11/99 (11%) | 03/51 (06%) | 1.9 (0.54-6.30) | 2.0 (0.52-7.40) | 0.79 (0.49-0.95) | 0.35 (0.27-0.43) | 0.70 (0.27-1.13) |
| **HIS criteria =2\*** | (ferritin + N/L ratio) | 35/96 (36%) | 11/51 (22%) | 1.70 (0.92-3.0) | 2.0 (0.93-4.50) | 0.76 (0.61-0.87) | 0.39 (0.29-0.49) |  |
| **Renal inflammation\*\*** | (Urea ≥ 70 mg/dL)<br>+Creatinine ≥ 1.5 mg/dL) | 33/93 (35%) | 14 /48 (29%) | 1.1 (0.69-1.90) | 1.2 (0.58-2.50) | 0.69 (0.54-0.81) | 0.35 (0.26-0.46) | 0.65 (0.48-0.81) |
| **Hyperglycemia\*\*\*** | (RBG ≥ 200 mg/dL) | 25/66 (38%) | 12/29 (41%) | 0.92 (0.54-1.60) | 0.86 (0.35-2.1) | 0.68 (0.50-0.82) | 0.29 (0.18-0.43) | 0.62 (0.43-0.80) |

\*In this bivariate analysis, presence of two criteria (hyperferritinaemia and hematological dysfunction) was analyzed. A total of 35 out of 96 (36%) male, and 11 out of 50 (22%) female COVID-19 patients developed both the HIC.
\*\*13 male and 8 female COVID-19 patients had chronic renal disease. Thus, (33-13= 20) male and 14-8 =6) female COVID-19 patients developed renal inflammation after SARS-CoV-2 infection.
\*\*\*11 male and 6 female COVID-19 patients had confirmed type -2 diabetic hyperglycemia, rest of the patients developed hyperglycemia after SARS-CoV-2 infection.

### Analysis of hyperinflammatory criteria

#### Hyperferritinaemia

Hyperferritinaemia (elevated serum ferritin) is considered as an independent risk factor associated with severity and mortality in COVID-19 (12). Hyperferritinaemia was present in 56% male and 30% female COVID-19 patients; the odd ratio was 2.9 (95% CI 1.4-6.0) with sensitivity 0.79 (95% CI 0.67 - 0.87). This suggests that the risk of male COVID-19 patients developing hyperferritinaemia is nearly three times the risk of female COVID-19 patients possess. Not only the proportion, mean and median values of serum ferritin were also much higher in male COVID-19 patients compared to their female counter parts (mean ferritin - 787 μg/L vs 473 μg/L; P <0.001). Since hyperferritinaemia suggests macrophage activation (4,12), the above observations suggest that SARS-CoV-2 infection leads to more severe inflammatory reactions in male compared to the female COVID-19 patients.

Although several blood parameters have been found altered in COVID-19 patients, N/L ratio has been used as a good indicator of hematological dysfunctions. Nearly 71% male, and 54% female COVID-19 patients had N/L ratio ≥10. The odd ratio was 2.1 (95% CI 1.0-4.2) with a sensitivity of 0.72 (95% CI 0.61-0.80) suggesting that the risk of male COVID-19 patients developing hematological dysfunctions is two times the risk female patients have. Also the mean and median values of N/L ratio were significantly higher in male compared to the female (16.3 vs. 12.5, P=0.009, table 1) COVID-19 patients suggesting more severe alterations in blood parameters, which could be due to a combined effect of neutrophilia and lymphocytopenia (13).

Lymphocytopenia was the most prominent feature of hematological dysfunctions; however, sharp differences were observed between male and female COVID-19 patients (Table 1). Approximately 86% male compared to only 64% female COVID-19 patients showed lymphocytopenia. These findings are in agreement with other studies in which development of sever lymphocytopenia in COVID-19 patients have been reported (13). However, it appears that SARS-CoV-2 induces more severe effect in male compared to female COVID-19 patients.

#### Cytokinaemia

COVID-19 has been suggested to induce severe cytokine storm in which several inflammatory cytokines or chemokines have been found elevated in COVID-19 patients (13). Among these are: interleukins, TNF-α, interferons, and CRP (2-4). However, CRP has been used recently for predicting the risk of severity and mortality in COVID-19 patients in a retrospective cohort study (11). Table 1 shows CRP values, which varied from 3 to 32 mg/dL with a median value of 5.5 mg/dL in male and 6.0 mg/dL in female COVID-19 patients. However, only 14% male and 12% female COVID-19 patients developed the criteria of cytokinaemia (CRP value ≥ 15 mg/dL) (Table 2). While the difference in CRP level between male and female COVID-19 patients wasn’t big as the odd ratio was 1.2 (95% CI 0.44 - 3.40). Despite the low percentage of male and female COVID-19 patents developing cytokinaemia, it was strongly associated with mortality in both sexes as discussed later.

#### Coagulopathy

A number of studies have reported increased D-dimer formation in COVID-19 patients and the risk is higher in patients with other co-morbidities suggesting coagulopathy as a potential pathological feature in COVID-19 patients (4,11). We randomly measured D-dimer formation in ∼40% patients and the mean values and the values associated with HIS are shown in table 1. Using the cut-off value of ≥ 1.0 μg/ml, we found 65% male and 55% female COVID-19 patients developed the criteria of coagulopathy (Table 1). The odd ratio was 1.50 (95% CI 0.50 - 4.60) suggesting that the risk male COVID-19 patients have for developing coagulopathy is 1.5 times the risk of female COVID-19 patients (Table 2).

#### Liver inflammation

Plasma AST activity was measured for assessing hepatic inflammation. As shown in table 1, the values of AST activity were slightly higher in male COVID-19 patients compared to the female patients but were not significant (P=0.067). Approximately 11% male and 6% female COVID-19 patients showed AST value ≥100 U/L; the cutoff limit for HIC. The odd ratio was 2.0 (95% CI 0.52 - 7.40), which suggests that male COVID-19 patients had twice the risk for developing liver inflammation than the female COVID-19 patients.

#### Renal inflammation

Several lines of evidence suggest that kidney dysfunctions could be early manifestation in COVID-19 patients and may substantially increase the risk for mortality (6,7,14). We also found several patients, with no previous history of renal complications, had elevated blood urea and serum creatinine levels suggesting possible pathogenic effect of SARS-CoV-2 infection on renal functions. Table 1 shows threshold values of blood urea and creatinine in male and female COVID-19 patients. We set the cut-off limits of blood urea as ≥ 70 mg/dL and creatinine as ≥ 1.5 mg/dL to establish the criteria of renal inflammation. As shown in table 2, 35% male and 29% female COVID-19 patients had blood urea urea ≥ 70 mg/dL (odd ratio: 1.2, 95% CI 0.58 - 2.50), whereas, 29% male and 19% female COVID-19 patients had creatinine level ≥ 1.5 mg/dL (odd ratio: 1.4, 95% CI 0.60 - 3.40). Moreover, 13% male and 16% female COVID-19 patients had chronic renal disease. Excluding these patients from the above population suggests that a significant proportion of COVID-19 patients developed renal inflammation after catching the disease. This suggests that SARS-CoV-2 infection induces severe renal inflammation in a significant proportion of both male and female COVID-19 patients.

A number of clinical studies have found that SARS-CoV-2 infection induces massive hyperglycemia, which may increase the severity of the diseases and risk for mortality (15-17). We measured RBG for assessing hyperglycemia and its association with mortality both in male and female COVID-19 patients and the values are shown in table 1. For establishing hyperglycemia, we set the cut-off value of RBG at ≥ 200 mg/dL. As shown in table 2, 38% male, and 41% female COVID-19 patients had RGB value ≥ 200 mg/dL. The odd ratio was 0.86 (95% CI 0.35 - 2.1), which suggests that female COVID-19 patients were more susceptible for developing hyperglycemia than the male counterparts. In addition, 11% male and 12% female had type-2 diabetes.

#### Association of various HIC with mortality

A detail analysis of the association of various HIC with mortality is presented in table 3. A total of 20 out of 100 male and 12 out of 51 female COVID-19 patients perished as of November 10, 2020. Among various HIS criteria, hyperferritinaemia (odd ratio: 1.33), cytokinaemia (odd ratio: 1.60), and hepatic inflammation (odd ratio: 1.30) were strongly associated with the mortality in male COVID-19 patients, whereas, coagulopathy (odd ratio: 3.3), and hematological dysfunctions (odd ratio: 1.70) were more strongly associated with the mortality in female COVID-19 patients.

**Table 3.** Association of various HIC with mortality in male and female COVID-19 patients

| HIC ↓ | Male<br>(N=20) | Female<br>(N=12) | Risk ratio (95% CI) | Odds ratio (95% CI) | Sensitivity (95% CI) |
| --- | --- | --- | --- | --- | --- |
| **Hyperferritinaemia** | 14/20 | 07/12 | 1.11 (0.63 - 2.00) | 1.70 (0.31 - 5.77) | 0.65 (0.41 - 0.85) |
| **Hematological dysfunction** | 15/20 | **10/12** | 0.90 (0.63 - 1.30) | **0.60 (0.10 - 3.70)** | 0.60 (0.39 - 0.79) |
| **Cytokinaemia** | 09/20 | 04/12 | 1.40 (0.53 - 3.40) | 1.60 (0.37 - 7.3) | 0.69 (0.39 - 0.91) |
| **Coagulopathy** | 11/20 | **09/12** | 0.81 (0.56 - 1.20) | **0.31 (0.03 - 3.20)** | 0.55 (0.33 - 0.77) |
| **Hepatic inflammation** | 04/20 | 02/12 | 1.2 (0.26 - 5.60) | 1.30 (0.19 - 8.10) | 0.67 (0.22 - 0.96) |
| **HIS criteria ≥ 2\*** | 16/20 | 10/12 | 0.96 (0.69 - 1.30) | 0.80 (0.12 - 5.20) | 0.62 (0.41 - 0.80) |
| **Renal inflammation\*\*** | 09/20 | 06/12 | 0.90 (0.43 - 1.90) | 0.82 (0.19 - 3.40) | 0.60 (0.32 - 0.84) |
| **Hyperglycemia** | 11/20 | 06/12 | 1.10 (0.55 - 2.20) | 1.20 (0.29 - 5.10) | 0.65 (0.38 - 0.86) |

\*Number of patients who perished having any two or more HIC excluding renal inflammation and hyperglycemia. As can be seen from the mortality data, 80% male and female COVID-19 patients died due to the development of two or more HIC.
\*\*6 out of 9 male and 5 out of 6 female COVID-19 patients 5 had chronic renal disease, suggesting severe impact of renal diseases on COVID-19 associated mortality.

In a bivariate analysis, influence of hyperferritinaemia and hematological dysfunction (N/L ratio) together was tested on mortality in male and female COVID-19 patients. A total of 35 out of 96 (36%) male, and 11 out of 51 (22%) female COVID-19 patients developed both these criteria of HIS. Out of these 9 male and 5 female COVID-19 patients died. This suggests that while less number of female COVID-19 patients developed hyperferritinaemia and hematological dysfunction together, they perished in very high proportion (5/12 = 45%) compared to the male COVID-19 patients (9/35 = 26%). Mortality proportion was increased further in female COVID-19 patients with chronic renal disease and/or diabetes. For instance, 5 out of 12 female and 6 out of 20 male COVID-19 patients, who died, had chronic renal disease. Similarly, 4 out of 12 female and 4 out of 20 male COVID-19 patients, who died, had type-2 diabetes. Thus, females with established renal pathology and /or diabetes carry much higher risk for mortality than males following SARS-CoV-2 infection.

#### Overall mortality

Figure 1 shows the trend of COVID-19 distribution (panel A) and mortality proportion (panel B) in male and female COVID-19 patients of different age groups. Regarding overall mortality, while it stayed at 5.30% (181/2997), the odd ratio for male to female mortality was 1.30 (95% CI 0.92 - 1.90) suggesting slightly higher risk for mortality in male COVID-19 patients. A total of 45 deaths out of 967 female COVID-19 patients and 114 deaths out of total 2030 male COVID-19 patients were recorded. The overall low mortality proportion that we observed in study, is in agreement to that has been observed in other parts/states of India (10). Most of the deaths were recorded in patients with age >40 years (Figure B) despite nearly 40% of the total COVID-19 cases were distributed in patients with age < 40 years (Figure 1 A). This suggests that young Indian male and female population is not at risk of SARS-CoV-2 associated mortality.

**Figure 1.**
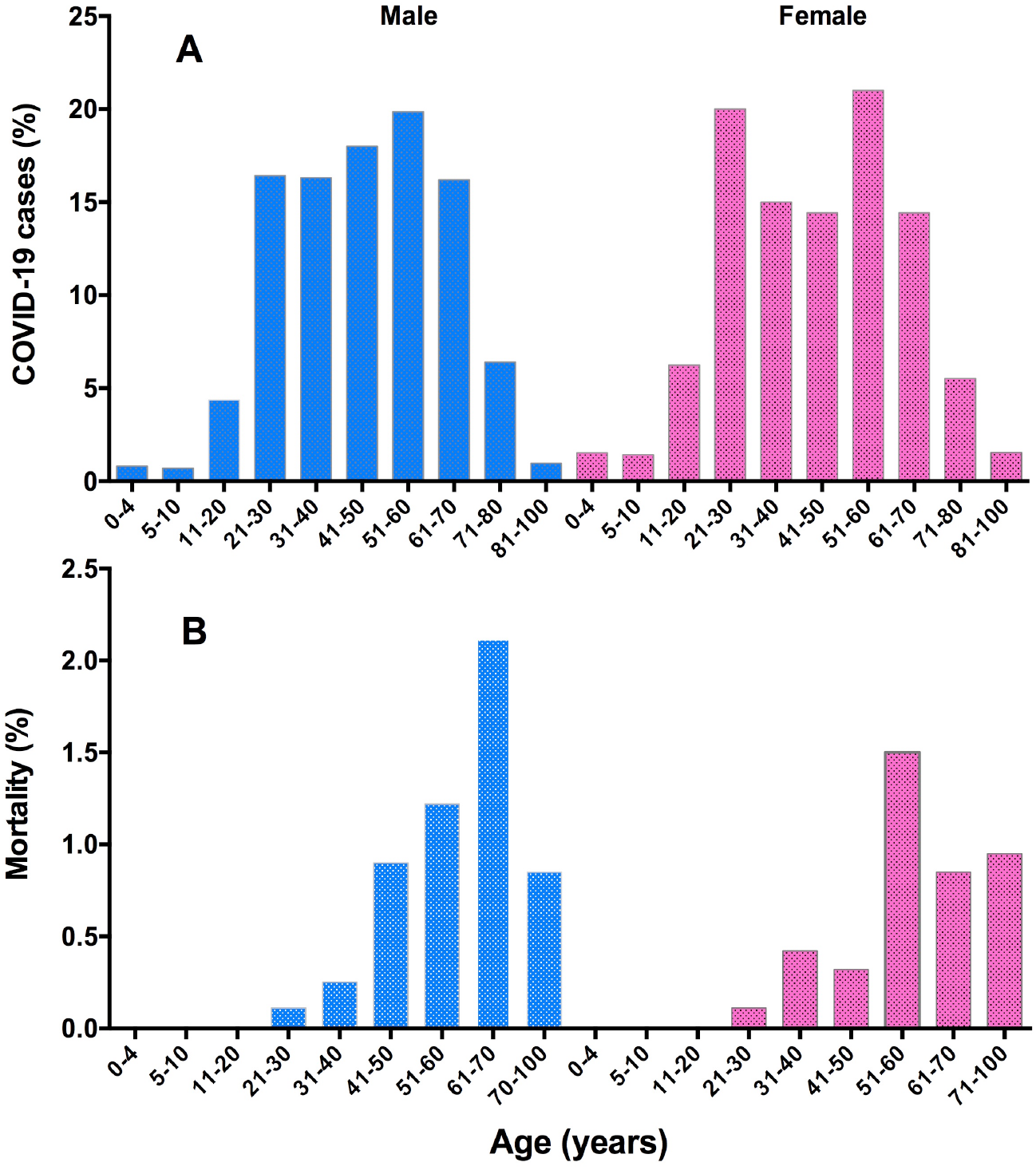
Trend of COVID-19 distribution (A), and mortality proportion (B), in male and female patients of different age groups. A total of 2997 COVID-19 cases (2030 males and 967 females) were included in this study.

Figure 2, shows Kaplan-Meier curve or survival curves of COVID-19 mortality. Most of the deaths were reported in older patients (age ≥50 years), and the survival curves of male and female COVID-19 patients were significantly not different (hazard ratio: 1.02, 95% CI 0.71-1.40, P = 0. 953), which suggest that older male and female population may have similar mortality hazard associated with COVID-19.

**Figure 2.**
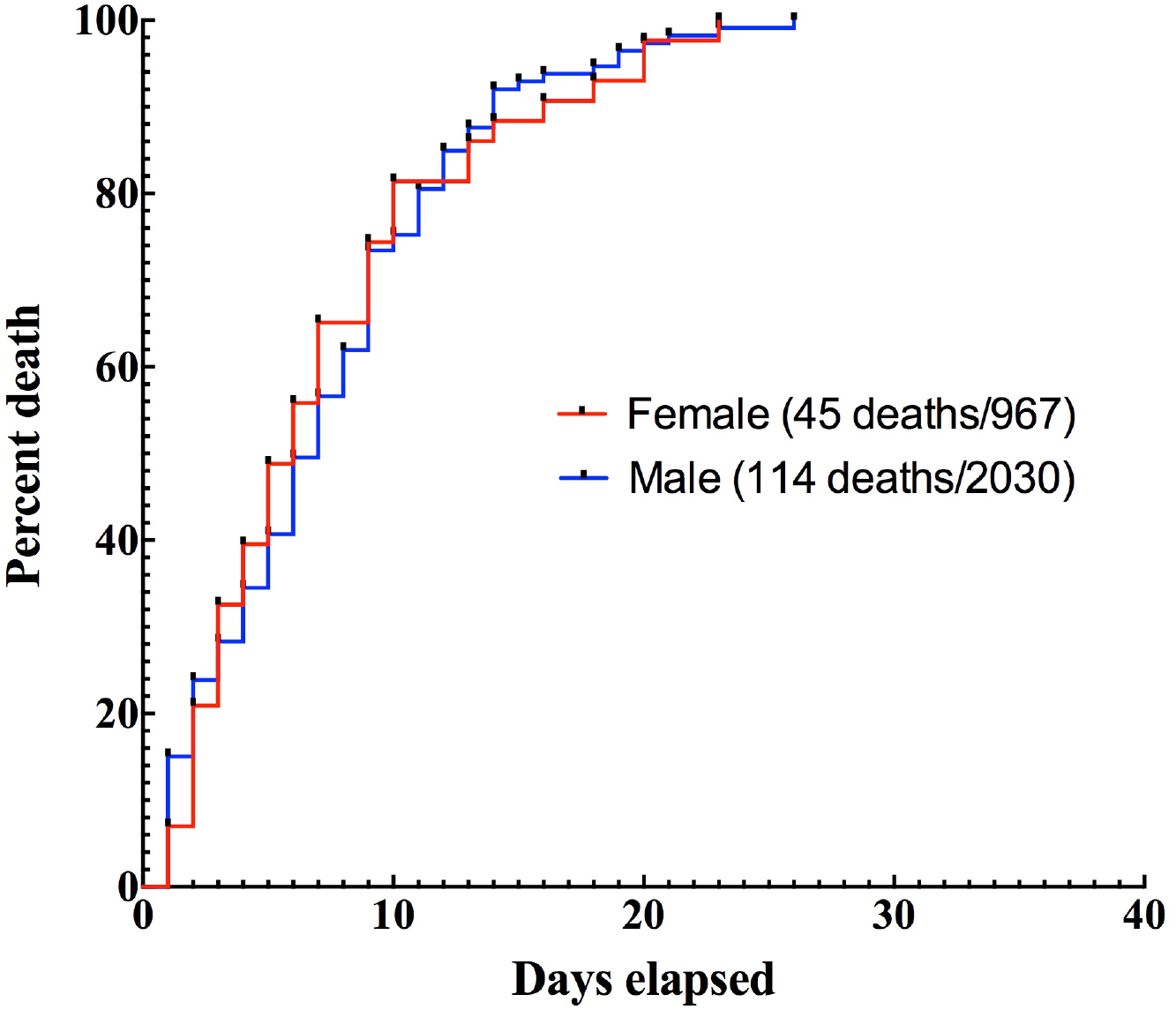
Kaplan-Meier curves/survival curves of COVID-19 patients who perished from among 2997 COVID-19 patients admitted between July 4 and December 3 2020. A total of 45 females and 114 males were expired. The survival curves of the two genders were not different (P = 0.869).

## Discussion

We have provided a comprehensive analysis for the development of various criteria of HIS and their association with mortality in the Indian COVID-19 patients. The results presented above suggest that male COVID-19 patients were more susceptible to develop hyperferritinaemia, hematological dysfunctions, hepatic inflammation, and coagulopathy compared to the female COVID-19 patients (Table 2). In a bivariate analysis also, male COVID-19 patients were two times more susceptible to develop hyperferritinaemia and hematological dysfunction than female COVID-19 patients (Table 2).

Regarding mortality, while hyperferritinaemia and cytokinaemia were strongly associated with mortality in male COVID-19 patients, coagulopathy, and hematological dysfunctions were more strongly associated with mortality in female COVID-19 patients.

Although nearly 30-35% male and female COVID-19 patients had renal inflammation, and ∼40% had hyperglycemia (Table 2), both these criteria were strongly associated with mortality in both male and female COVID-19 patients (Table 3). However, female patients with chronic renal disease and diabetes had more deaths than male COVID-19 patients. Despite these gender disparities and more males than females developed COVID-19, the survival curves or mortality hazard associated with male and female COVID-19 patients were not different suggesting that both male and female COVID-19 patients may die in similar proportion within the same duration of time.

A recent study by Webb et al (2020) proposed that development of two or more criteria of hyperinflammatory syndrome leads to mechanical ventilation and death in COVID-19 patients (11). We also found that nearly 80% male and female COVID-19 patients, who died had developed two or more HIC. Thus, our findings provide experimental validation for the application of various criteria of in predicting the risk of severity and mortality in COVID-19 patients as proposed recently (11). The remaining patients had at least one HIC and either had hyperglycemia or severe renal injury or both as additional risk factor associated with mortality (Table 3). These findings are in agreement with other studies, which have reported that diabetes/hyperglycemia and renal injury/inflammation substantially increase the risk of mortality in COVID-19 patients (6-8,15,16).

Epidemiological data suggests that males carry higher risk than females for SARS-CoV-2 transmission (10,11). Our data also supports these findings because, in a total of 2997 COVID-19 patients admitted to ELMCH between July 4 to December 3 2020, male to female odd ratio was 2.0 suggesting that males carry two times more risk than female for SARS-CoV-2 infection. The reason for this is not clear; however, a potent lymphocyte and monocyte network may act as a potential barrier, thereby, restricting virus propagation in females (4,18,19). In support of this, we found that the mean/average value of lymphocyte count was significantly higher in female compared to the male COVID-19 patients (Table 1). About 86% male compared to 64% female patients showed lymphocytopenia (P=0.012). More importantly, when we analyzed Pearson product moment correlation, we found strong negative correlation between serum ferritin and both lymphocyte and monocyte counts in female but not male COVID-19 patients. This simply suggests that higher lymphocyte and monocyte counts may have negative or suppressive effect on ferritin elevation (macrophage activation) in female but not in male COVID-19 patients. These findings are in agreement with earlier reports, which suggest that females possess stronger immunity than males, which probably provides them protection against SARS-CoV-2 infection. Indeed previous studies have shown that females of the age ≥ 50 years display higher lymphocyte count and lower neutrophil to lymphocyte ratio than the males of the same age (19-21). In addition, some reports suggest that circulating estrogens and progesterone in females may also provide protection against SARS-CoV-2 infection (22).

Altogether, the findings discussed in this study suggest that male COVID-19 patients develop more severe HIC than female patients, and distinct HIC may be associated with mortality in male and female COVID-19 patients. Moreover, female COVID-19 patients with chronic renal pathology and diabetes carry severely elevated risk for mortality than male COVID-19 patients. Thus, gender differences associated with COVID-19 severity and mortality, which have been reported in various epidemiological studies, may arise due to differences in various HIC. These results may help in developing personalized or gender based treatments for COVID-19 patients.

## Data Availability

All the data is available with the corresponding author

## Acknowledgments

Facilities provided by the department of Personalized and Molecular Medicine and the Vice Chancellor’s office, ERA University are gratefully acknowledged. All authors are cordially thankful to the medical and paramedical staff of Era’s Lucknow Medical College and Hospitals for their selfless care and treatment provided to the COVID-19 patients and help in completing this study.

## Ethics approval

Ethical Committee of ERA University, and ELMCH approved the study protocol

## Consent to participate

Written consent from participants to conduct and publish this study was taken

## Availability of data and material

All the data is available with the corresponding author

## Authors’ contributions

All authors have contributed and approved this study

